# Non-Linear Size Effects and Episodic Progression in the Ascending Aorta

**DOI:** 10.1101/2025.09.16.25335949

**Authors:** Carlos Alberto Campello Jorge, Prabhvir S. Marway, Gregory Spahlinger, Heather A. Knauer, Marion Hofmann-Bowman, Venkatesh L. Murthy, Nicholas S. Burris

**Author notes:** <u>Address for correspondence:</u> Nicholas S. Burris, M.D., Department of Radiology, University of Wisconsin-Madison, 800 University Bay Drive, Suite 210, Rm 224, Madison, WI 53705. <u>Funding:</u> Nicholas S. Burris - National Institutes of Health (R01HL170059).

## Abstract

**Background:** Predicting ascending aortic (AsAo) growth is challenging. Conventional paradigms often assume a linear and monotonic relationship between baseline size and future growth that occurs continuously–assumptions that may oversimplify biology-driven disease progression. We first evaluated whether body size–indexed baseline AsAo diameter shows a non-linear association with subsequent growth at the population-level, and second whether patient-level growth trajectories are predominantly continuous or episodic.

**Methods:** We performed a single-center, retrospective study (2012–2024). The Primary Cohort (n=3,315; ≥2 CT/MR scans) was used to model the relationship between baseline indexed AsAo size (Z-score) and subsequent annualized growth using multivariable linear regression and generalized additive models (GAMs), adjusting for clinical covariates. A Sub-Cohort (n=1,055; ≥4 scans) was used to classify longitudinal phenotypes as: Stable (Total Growth <2.0 mm), Stable-with-Noise (Total Growth <2.0 mm with alternating small changes), Continuous Growth (Total Growth ≥2.0 mm without a qualifying event), or Discontinuous/Episodic Growth (Total Growth ≥2.0 mm with ≥1 “growth event”). A growth event was defined as a diameter increase ≥2.0 mm within a single imaging interval or across two adjacent intervals (combined 0.5–5 years).

**Results:** In the Primary Cohort, baseline Z-score demonstrated a significant non-linear (U-shaped) association with subsequent growth in the GAM (p<0.001), with higher growth at both small (Z<0) and severely dilated (Z>5) sizes. In the Sub-Cohort examining growth trajectory, the distribution was: Stable: 50.4% (532/1,055), Stable-with-Noise: 21.6% (228/1,055), Continuous: 5.4% (57/1,055), and Discontinuous/Episodic 22.6% (238/1,055). Among patients with measurable growth (Total Growth ≥2.0 mm, n=295), 81%(n=238) exhibited episodic growth and 58.3% (172/295) had a non-dilated baseline aorta (Z<2). Group differences (e.g., younger age and smaller baseline Z-scores in the Growth vs. Stable groups) were consistent across sensitivity analyses.

**Conclusions:** AsAo growth is not well described by linear, continuous assumptions. Baseline size relates to future growth in a non-linear (U-shaped) manner, and nearly one-quarter of patients exhibit discrete growth bursts separated by periods of quiescence, with episodic behavior dominating among those who enlarge. These findings support a punctuated growth paradigm and argue for re-examining surveillance intervals, risk communication, and threshold-based decision pathways in thoracic aortic disease.

## INTRODUCTION

Predicting future ascending aortic (AsAo) growth and the risk of life-threatening complications (e.g., type A dissection) remains a central challenge in cardiovascular medicine. Current clinical management is heavily influenced by maximal aortic diameter, and to a lesser degree the rate of growth as a maker of disease progression.^1,2^ The majority of research studies, risk models and clinical intuition are based on an assumption of linear growth, where the overall rate of change is between imaging studies assumed to be relatively consistent over time. While convenient, this framework has two potential major flaws.

First, it presumes a nearly linear relationship between aortic size and future growth rate, as has been suggested in prior studies using regression-based models.^3-5^ However, such an assumption may be biologically implausible, as the aortic wall is not a passive structure but rather a dynamic, biologically active tissue capable of remodeling its composition in response to various stresses. While processes such as smooth muscle cell phenotypic switching^6^ and extracellular matrix turnover^7^ may promote disease progression, stress-mediated wall remodeling^8^ may act as a compensatory mechanism that blunts growth – at least until this capacity is exhausted, at which point accelerated dilation may occur. This dynamic interplay makes it unlikely that disease progression follows a uniform, continuous trajectory; instead, growth may follow curves more typical of compensated versus decompensated biological processes (e.g., Gompertz function). Supporting this concept, recent studies have demonstrated little to no association between baseline aortic size and subsequent growth rate^9^, and continue to highlight the paradoxical relationship between an aortic size and an individual patient’s risk – over 60% of fatal type A dissections occur below commonly accepted dilation (<4.0 cm).^10^

Second, beyond the relationship between baseline size and growth rate, the temporal pattern of aortic enlargement within an individual may also be non-linear. Clinically, many patients demonstrate long periods of quiescence, with little to no change in aortic dimensions, punctuated by episodes of accelerated growth – a phenomenon that has been suggested in abdominal aortic aneurysm.^11,12^ These fluctuations are biologically plausible given the interplay between disease-promoting processes and finite compensatory mechanisms that stabilize wall integrity. Despite this, the most common clinical approach is to estimate growth by comparing diameters between two time points—often a remote baseline and a recent follow-up—to reduce the impact of measurement variability. This method inherently implies a linear growth trajectory and therefore captures only an ‘average’ growth rate. Such an approach may obscure underlying episodic growth dynamics, potentially overlooking critical periods of disease activity and heightened risk.

This study had two primary objectives. First, to characterize the relationship between initial ascending aortic size and subsequent growth rate in a large surveillance cohort, using flexible statistical models capable of depicting potential non-linear associations. Second, to examine the temporal growth pattern in a subgroup of patients with long-term serial imaging, to determine whether aortic enlargement occurs as a steady, continuous process or in episodic bursts. We hypothesized that the conventional linear-continuous paradigm oversimplifies the biology of aortic disease, and that ascending aortic growth is more accurately described by non-linear and episodic dynamics.

## METHODS

### Study Design and Population

This single-center, retrospective cohort study was conducted on data collected between January 2000 and December 2023, after approval from the local institutional review board, with waiver of informed consent. The study was reported in accordance with the Strengthening the Reporting of Observational Studies in Epidemiology (STROBE) guidelines. Patients were identified from existing institutional aortic research databases and through searches of the electronic medical record. We included consecutive adult patients (≥18 years) who had at least two CT or MR angiograms of the thoracic aorta with a minimum surveillance interval of six months. Patients with any prior history of aortic repair or aortic dissection were excluded. Electronic medical records were used to collect baseline demographics, comorbidities (e.g., hypertension, diabetes mellitus, smoking history, known heritable thoracic aortic disease and confirmed chart diagnosis of aortitis), vital signs (e.g., blood pressure), medications, and clinical outcomes (e.g., type A aortic dissection, prophylactic aortic repair, aorta-specific mortality).

Our analyses were conducted on two distinct cohorts derived from this population. The **Primary Cohort**, comprising all eligible patients, was used for a *population-level* analysis of the association between baseline aortic diameter and growth rate. From this group, a “Growth Trajectory” **Sub-Cohort** of patients with at least four serial CT or MR angiograms, was used for a detailed *patient-level analysis* of growth trajectories and patterns.

### Aortic Measurements and Classification of Growth Patterns

Aortic diameter measurements were extracted from structured measurement subsections of clinical radiologic reports using an in-house text processing pipeline. All aortic measurements were extracted from clinical radiologic reports, which at our center are performed in a dedicated 3D imaging laboratory using standardized protocols with cardiothoracic radiologist oversight. To ensure consistency of measurement location across all cases and time-points, the mid-ascending aorta (mid-AsAo) landmark was used to extract diameter measurements for analysis, with manual review performed in cases with suspected typographical errors due to implausible values, which were corrected where identified (e.g., interval change >10 mm, absolute size >70mm or <20mm). To account for known variability in aortic size with patient factors such as body size (i.e., body surface area), age and sex, all diameter measurements in this study were transformed to z-scores using a common nomogram for aortic size in adults.^13^

From these longitudinal measurements, we defined two key metrics to classify patient-level growth dynamics within the Growth Trajectory Sub-Cohort. First, Total Growth was calculated for each patient as the overall change in diameter predicted by a linear regression model fitted to all available measurements, a method that minimizes the impact of single-interval measurement variability. Second, an episodic growth event was defined as a diameter increase of ≥2.0 mm occurring either within a single imaging interval or across two adjacent intervals with a combined duration of 0.5-5 years. This threshold was selected given that this magnitude of change exceeds typical inter-scan measurement variability.^14,15^

Using these two metrics, patients in the Growth Trajectory Sub-Cohort were further categorized into four mutually exclusive growth patterns. Patients were initially stratified by their overall growth into Stable (Total Growth <2 mm) or Growth (Total Growth ≥2 mm) groups. Each of these groups was then further stratified by the presence or absence of an episodic growth event. This yielded the final four analysis groups: 1) Stable – negligible overall growth (<2 mm Total Growth and no episodic events); 2) Stable with Noise – negligible overall growth (<2 mm Total Growth) with at least one interval meeting the criteria for a growth episode, likely reflecting measurement variability in most cases; 3) Continuous Growth – significant overall growth (≥2.0 mm Total Growth) without any single interval meeting the criteria for an episodic event; and 4) Episodic Growth – significant overall growth (≥2.0 mm Total Growth) with at least one episodic event (≥2.0 mm Total Growth).

### Statistical Analysis

All statistical analyses were performed using Python (Version 3.13.2 using the pandas, numpy, matplotlib, seaborn, scipy packages) and R software (Version 4.4.0 using the tidyverse, emmeans, mgcv packages).^16^ Continuous variables were summarized as mean ± standard deviation or median and interquartile range (IQR), while categorical variables were presented as frequencies and percentages, with a two-sided p-value < 0.05 considered statistically significant. For the population-level analysis on the Primary Cohort, we employed a hierarchical modeling approach to comprehensively evaluate the relationship between baseline size and growth rate. We first assessed the conventional linear assumption by sequentially fitting a simple linear regression model, a multivariable model adjusting for clinical covariates, and a final multivariable model including pre-specified interaction terms. Furthermore, to test the hypothesis that the relationship between baseline size and growth rate changes at the clinical threshold for aortic dilation, we fitted a piecewise linear regression model. A single knot was pre-specified at a *Z-*score of 1.96, allowing the slope to differ for normal versus dilated aortas. To explore more complex, non-linear relationships, we then fitted a parallel series of Generalized Additive Models (GAMs) using penalized thin plate regression splines to model the non-linear effect of baseline aortic size (*Z-*score), again progressing from a simple model to a multivariable model and a final multivariable model with interaction effects. For both linear and GAM analyses, the added contribution of each interaction term was quantified by comparing the full interaction model to a main-effects-only model and reporting the change in R-squared (ΔR^2^). The performance of all models was compared using the Akaike Information Criterion (AIC) to identify the best balance of model fit and complexity.

For the patient-level analysis of the Sub-Cohort, we compared baseline characteristics across the four predefined growth patterns using ANOVA, Kruskal-Wallis, or chi-squared tests as appropriate. We performed sensitivity analyses by repeating the primary analyses restricting the cohort to patients without known syndromic aortopathies.

## RESULTS

### Study population - Primary Cohort

Overall, we identified a Primary Cohort of 3,315 patients meeting inclusion criteria, with a median age of 62.4 years (IQR: 53.2–70.7), and a majority being male (68%, n=2,252) and with a history of hypertension (85%, n=2,820). Heritable thoracic aortic disease was present in a minority (3.4%, n=113), with Marfan syndrome being the most common form (2.5%, n=84). The median baseline mid-ascending aortic Z-score was 1.58 (IQR: 0.06–3.04), corresponding to a median absolute diameter of 40.4 mm (IQR: 36.0–44.0). Over a median imaging follow-up of 3.5 years (IQR: 1.6-6.4), the median annualized growth rate was 0.07 mm/year. A complete summary of the cohort characteristics is available in **Table 1**.

**Table 1:**
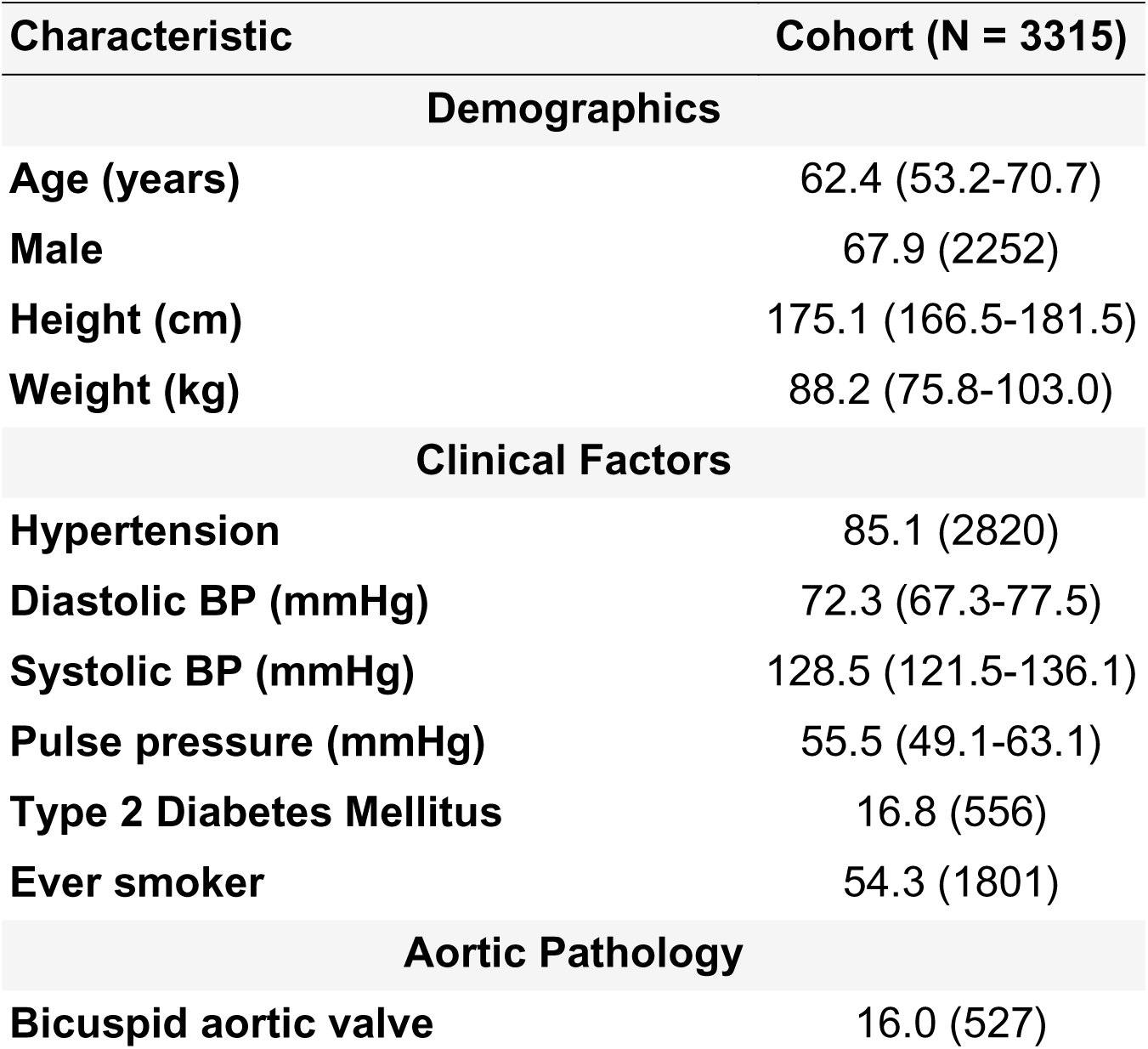
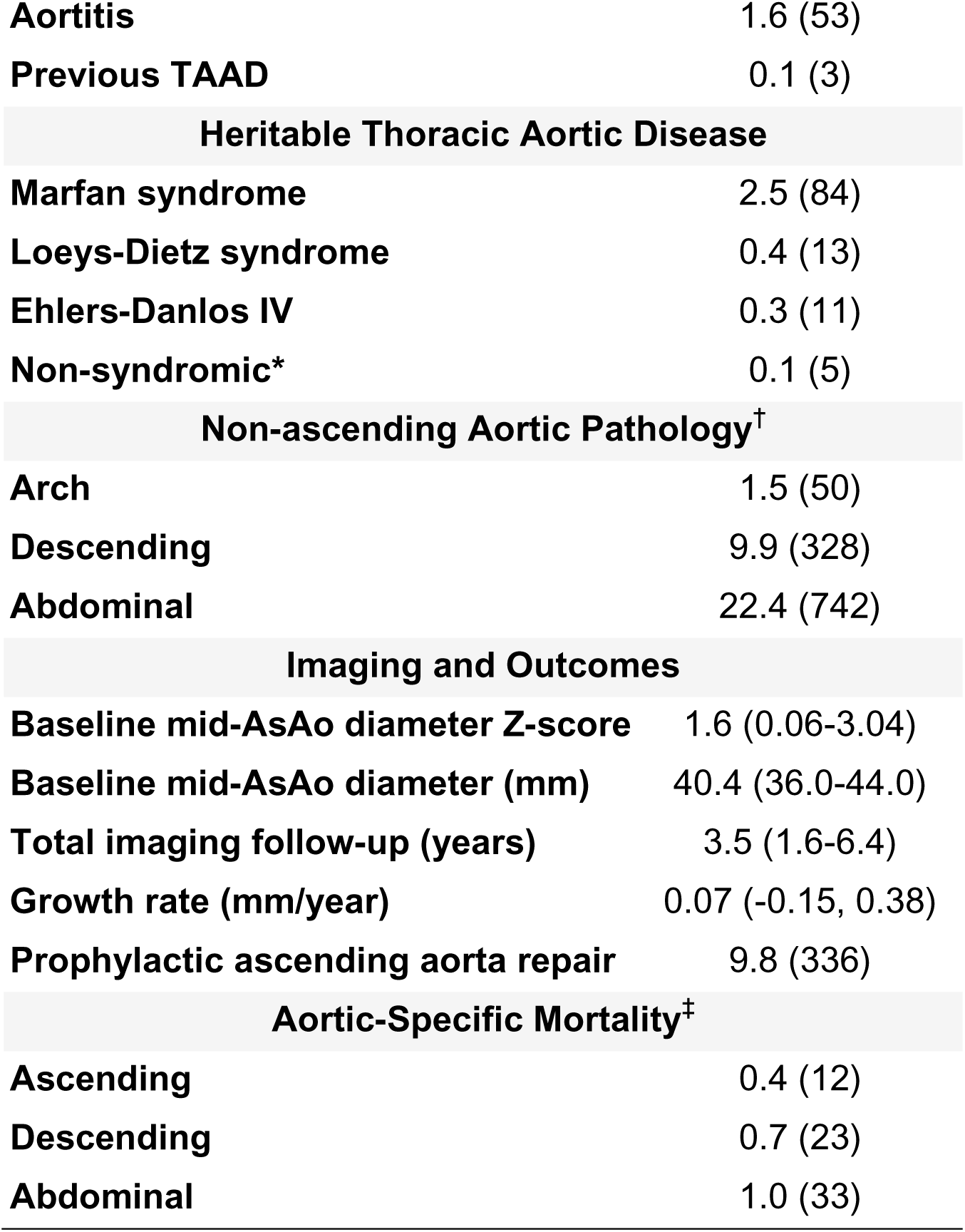
Baseline Demographics, Clinical Characteristics, and Outcomes of the Primary Cohort. Median (IRQ) or %(N) *includes ACTA2 (n=3), MYH11 (n=1), and MYLK (n=1) pathogenic variants. †Represents aneurysm, dissection, or open or endovascular repair in the non-ascending aorta. ‡Aortic diseases implicated (aneurysm, dissection, rupture, or operative intervention) in Part I of Death Certification. Data are presented as median (IQR) or % (n), unless otherwise specified. Abbreviations: AsAo, ascending aorta; BP, blood pressure; IQR, Interquartile Range; TAAD, Type A Aortic Dissection.

| Characteristic | Cohort (N = 3315) |
| --- | --- |
| **Demographics** |  |
| **Age (years)** | 62.4 (53.2-70.7) |
| **Male** | 67.9 (2252) |
| **Height (cm)** | 175.1 (166.5-181.5) |
| **Weight (kg)** | 88.2 (75.8-103.0) |
| **Clinical Factors** |  |
| **Hypertension** | 85.1 (2820) |
| **Diastolic BP (mmHg)** | 72.3 (67.3-77.5) |
| **Systolic BP (mmHg)** | 128.5 (121.5-136.1) |
| **Pulse pressure (mmHg)** | 55.5 (49.1-63.1) |
| **Type 2 Diabetes Mellitus** | 16.8 (556) |
| **Ever smoker** | 54.3 (1801) |
| **Aortic Pathology** |  |
| **Bicuspid aortic valve** | 16.0 (527) |
| **Aortitis** | 1.6 (53) |
| **Previous TAAD** | 0.1 (3) |
| **Heritable Thoracic Aortic Disease** |  |
| **Marfan syndrome** | 2.5 (84) |
| **Loeys-Dietz syndrome** | 0.4 (13) |
| **Ehlers-Danlos IV** | 0.3 (11) |
| **Non-syndromic\*** | 0.1 (5) |
| **Non-ascending Aortic Pathology<sup>†</sup>** |  |
| **Arch** | 1.5 (50) |
| **Descending** | 9.9 (328) |
| **Abdominal** | 22.4 (742) |
| **Imaging and Outcomes** |  |
| **Baseline mid-AsAo diameter Z-score** | 1.6 (0.06-3.04) |
| **Baseline mid-AsAo diameter (mm)** | 40.4 (36.0-44.0) |
| **Total imaging follow-up (years)** | 3.5 (1.6-6.4) |
| **Growth rate (mm/year)** | 0.07 (-0.15, 0.38) |
| **Prophylactic ascending aorta repair** | 9.8 (336) |
| **Aortic-Specific Mortality<sup>‡</sup>** |  |
| **Ascending** | 0.4 (12) |
| **Descending** | 0.7 (23) |
| **Abdominal** | 1.0 (33) |

Several clinical characteristics were found to be independent predictors of baseline aortic size (Z-score) by multivariable linear regression (R^2^ = 0.147). Specifically, older age and bicuspid aortic valve were associated with a higher Z-score. Conversely, male sex, higher pulse pressure, Type 2 Diabetes, aortitis, and Marfan syndrome were all associated with a lower Z-score (**Table S1**).

### Primary Cohort - Indexed Baseline Aortic Size versus Growth (Population-level)

#### Linear Model

Employing a simple linear model, baseline Z-score was not a significant predictor of growth rate (p=0.199, R^2^=0.001, **Figure 1**), and this observation persisted after adjusting for clinical covariates (p=0.319, adjusted R^2^=0.002). Adopting a piecewise linear model (knot at Z-score 2) revealed a significant change in slope (interaction p<0.001) and improved fit (ΔAIC= −35), inconsistent with a single linear relation (**Figure 2**).

**Figure 1.**
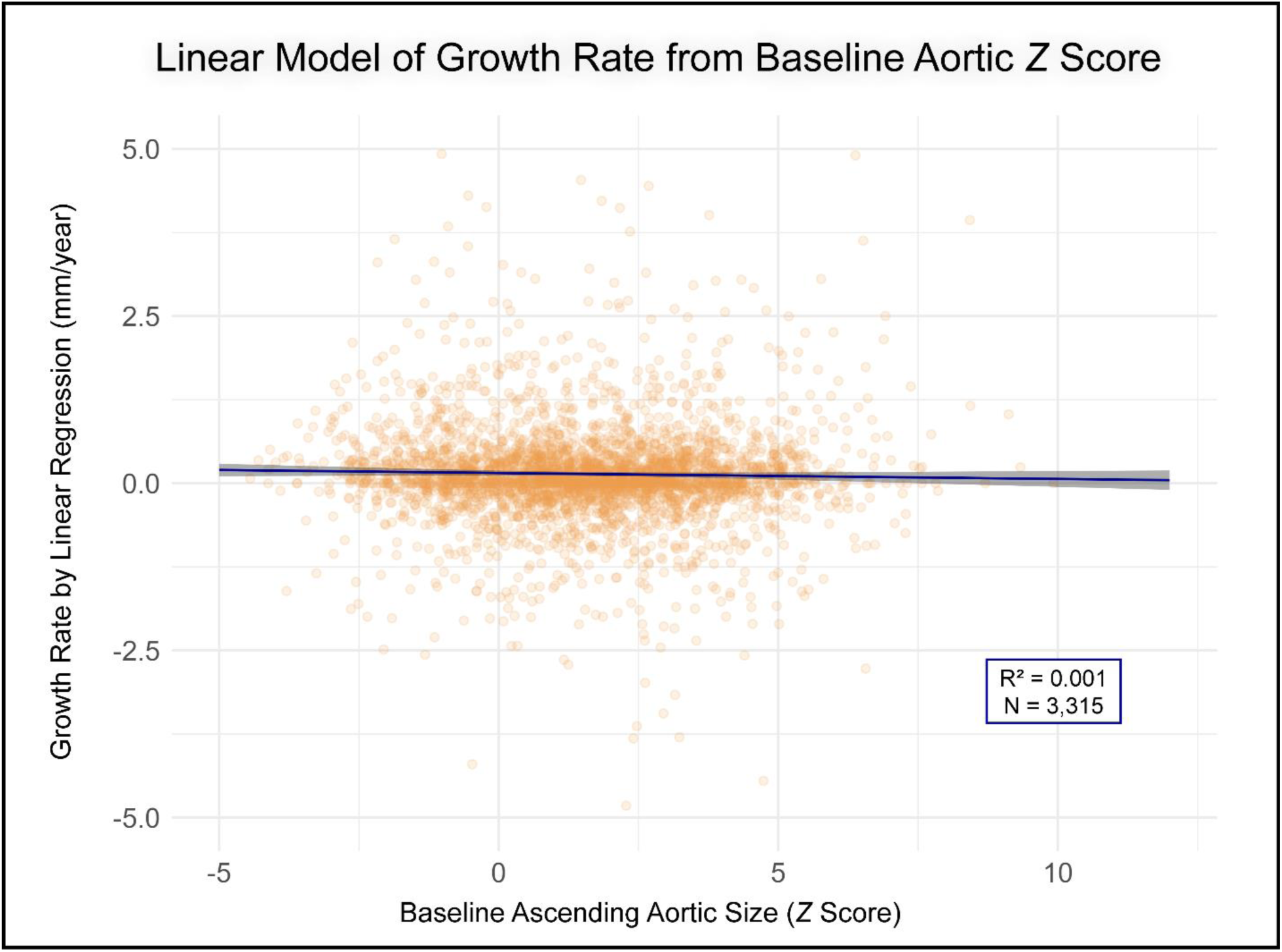
Relationship Between Baseline Aortic Z-Score and Growth Rate Modeled with Simple Linear Regression. This plot shows individual patient growth rates (orange points) against their initial Z-score, with the blue line representing the fitted linear regression model. The shaded gray area indicates the 95% confidence interval for the regression line. Plot axes are truncated for improved visualization outlier points (n=17) with an initial Z-score >12.5 or an absolute growth rate >5 mm/year not displayed.

**Figure 2.**
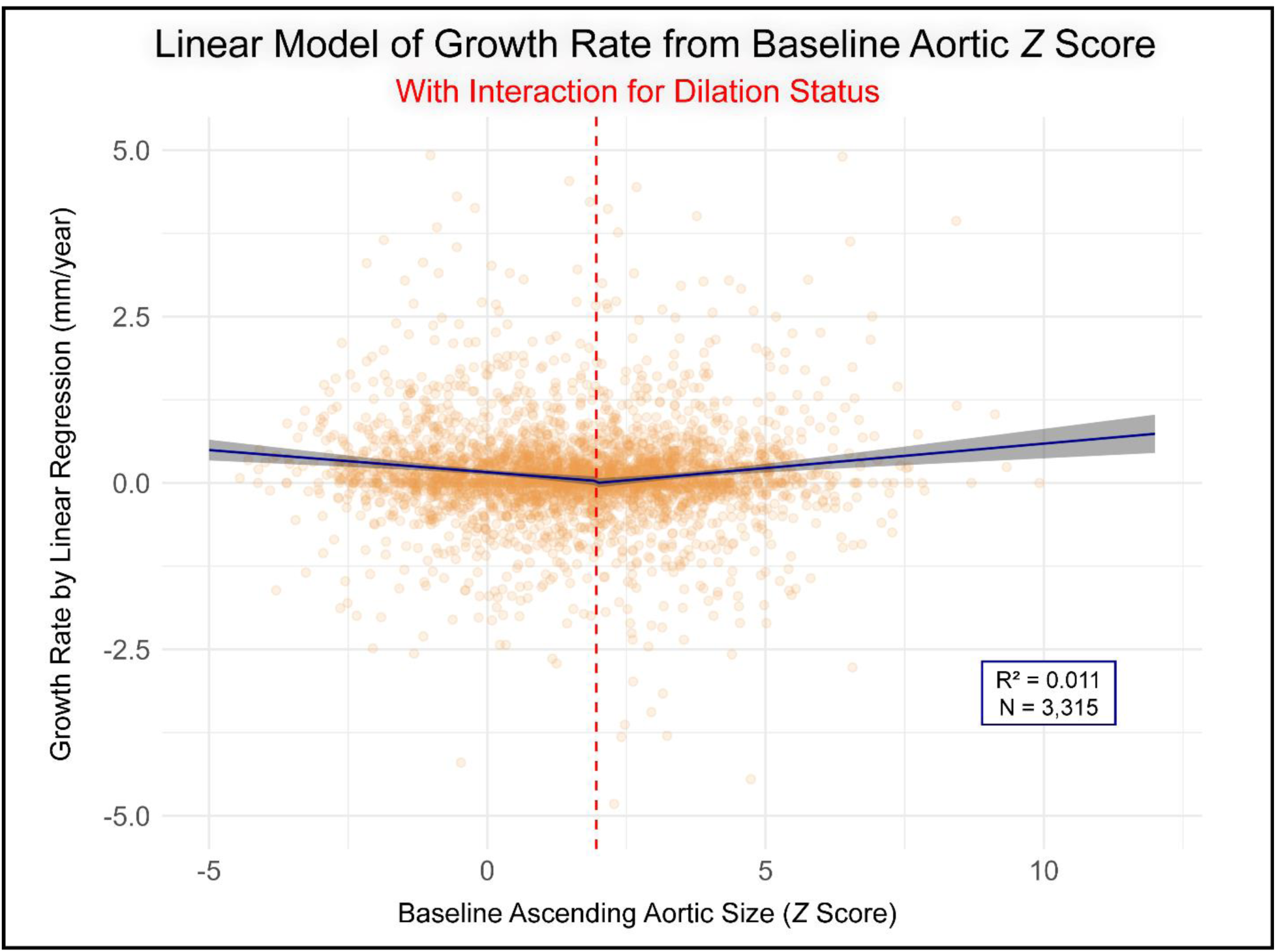
Relationship Between Baseline Aortic Z-Score and Growth Rate Modeled with a Linear Interaction for Dilation Status. This plot shows the fitted linear regression model (blue line) with its 95% confidence interval (shaded gray area) over the raw patient data (orange points). The model includes an interaction term at the dilation threshold (red dashed line), allowing the slope of the relationship between Z-score and growth rate to differ for patients with normal versus dilated aortas. Plot axes are truncated for improved visualization outlier points (n=17) with an initial Z-score >12.5 or an absolute growth rate >5 mm/year not displayed.

In a secondary analysis, the relationship between baseline aortic size and growth was significantly modified by age, sex, pulse pressure, and systolic blood pressure (all interaction p<0.01; ΔR² 0.003–0.013). A consistent pattern emerged in which the size– growth relationship inverted across clinically distinct subgroups. In younger patients (<50 years), smaller aortas grew faster, whereas larger aortas grew more slowly (slope −0.06 mm/yr per Z, p=0.004), while in older patients (80 years) the trend was weakly positive but not significant. In females, larger baseline size predicted faster growth (+0.01 mm/yr per mm, p=0.005), whereas in males the relationship was reversed, with larger size associated with slower growth (−0.02 mm/yr per mm, p<0.001). Similarly, at normal pressures (45 mmHg pulse pressure; 100 mmHg systolic blood pressure), smaller aortas tended to grow faster and larger aortas more slowly (−0.05 to −0.06 mm/yr per Z), but at elevated pressures (65 mmHg pulse pressure; 140 mmHg systolic blood pressure) the relationship inverted, with larger aortas trending toward faster growth (+0.01 to +0.02 mm/yr), though not consistently significant. Detailed results are provided in **Table S2** and visualized in **Figures S1-S4**.

#### Generalized Additive Model (GAM)

To further explore potential non-linear relationships between baseline aortic Z-score and growth rate, GAM was subsequently employed. By GAM we observed a significant, U-shaped relationship between baseline aortic Z-score and future growth rate (p<0.001; R^2^ = 0.017), as shown in **Figure 3 (Left Panel)**. This non-linear relationship was characterized by higher average growth rate in normal-sized aortas (Z< 0; diameter< ≈ 36 mm), followed by slower average growth in the mild-to-moderate dilation range (Z ≈ +2 to +5; diameter ≈ 36-48 mm) before demonstrating a clear inflection towards higher growth rate, after which the growth rate increased again in severely dilated aortas (Z>5; diameter > ≈49 mm), as shown in **Figure 3 (Right Panel)**. To further characterize the patient populations across this non-monotonic curve, we stratified the cohort at the apparent inflection points of Z=0 and Z=+5. Patients with severely dilated aortas (Z>5) were found to be significantly less likely to be male and had a higher prevalence of bicuspid aortic valve compared to those in the mild-to-moderate dilation group. Conversely, patients in the smallest ascending aortic size category (Z<0) were notably younger on average, had higher rates of genetic aortopathy (e.g., Marfan syndrome) as well as aortic disease affecting distal segments (i.e., arch, descending and abdominal aorta). These findings, as detailed in **Table S3**. After adjusting for the non-linear effect of baseline aortic size, no other clinical covariates, including age, sex, or Marfan syndrome, remained significant predictors of growth rate in the model (all p>0.05).

**Figure 3.**
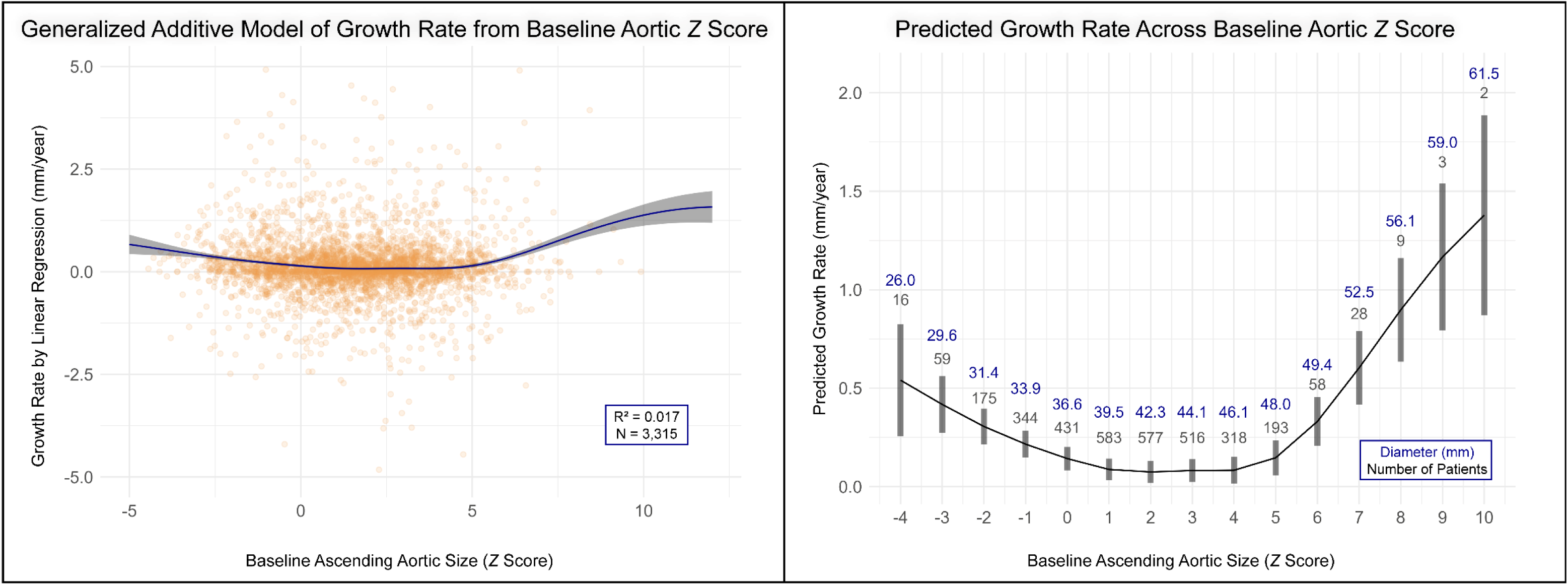
Non-Linear Relationship Between Baseline Aortic Size and Growth Rate. **(Left Panel)** The continuous relationship from a Generalized Additive Model is displayed, with the solid line representing the predicted mean growth rate and the shaded area indicating the 95% confidence interval, plotted over the individual patient data (scatter points). The relationship is significantly non-linear (p<0.001), forming a U-shaped curve with accelerated growth at both low (Z-score < 0) and high (Z-score > 5) initial sizes. For visualization, the plot axes have been truncated, excluding 17 outliers. **(Right Panel)** The same model results are shown as estimated marginal means for each integer Z-score bin. Gray bars represent the 95% confidence interval of the mean estimate. The patient count (N) and corresponding median aortic diameter (mm) are labeled above each bar.

Subsequent analysis revealed that several patient characteristics were significant effect modifiers, altering the relationship between aortic size and growth. We found significant non-linear interactions in the GAM between baseline Z-score and sex (p<0.001; deviance explained 2.5%, +0.8%), age (p<0.001; 3.6%, +1.9%), pulse pressure (p<0.001; 3.6%, +1.9%), and systolic blood pressure (p<0.001; 3.2%, +1.5%), as shown in **Figures 4 & S5–S7**. For patient age, a significant non-linear interaction was observed, particularly for Z-scores between +2 and +6, where the growth rate was notably faster in younger patients compared to older patients with similarly sized aortas (**Figure S5**). The size-growth relationship also differed meaningfully by sex: in women, growth was substantially faster at larger/aneurysmal sizes, while in men this interaction was blunted (**Figure 4**). The impact of systolic blood pressure was most pronounced in smaller, non-dilated aortas, where higher pressures were associated with accelerated growth (**Figure S6**). Conversely, the effect of pulse pressure became evident in severely dilated aortas (Z-score >5), where higher pulse pressure was linked to faster growth rates (**Figure S7**). Other patient factors, including Marfan syndrome, aortitis, and diabetes, did not significantly moderate the size-growth rate relationship (**Table S4**).

**Figure 4.**
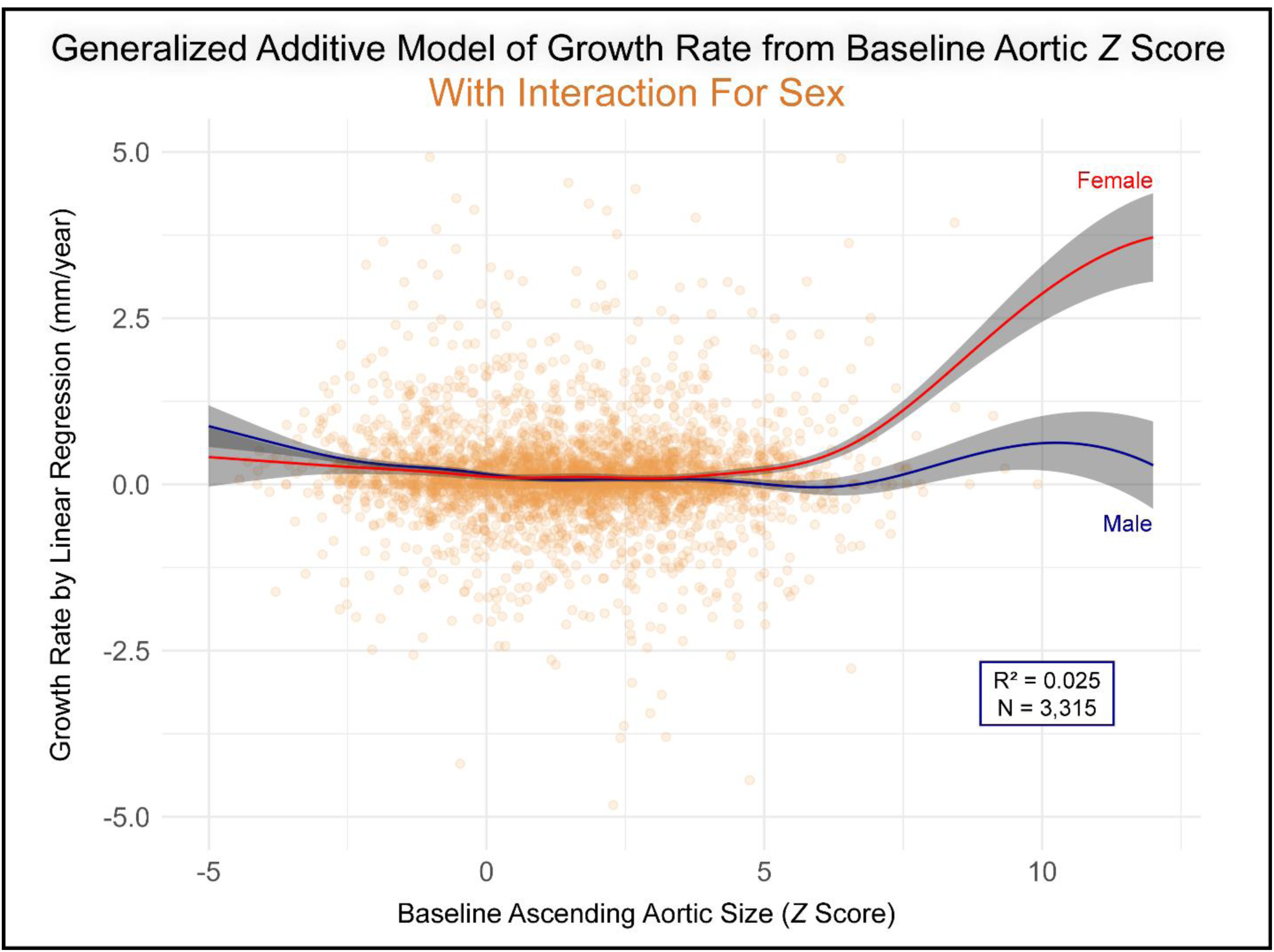
Non-Linear Interaction Between Baseline Aortic Z-Score and Sex on Aortic Growth Rate in a Generalized Additive Model. This plot visualizes the significant non-linear interaction between baseline Z-score and sex from the Generalized Additive Model. Smooth curves, representing the predicted growth rates, are shown for females (red) and males (blue), with their respective 95% confidence intervals (shaded gray areas) plotted over the raw patient data (orange points). The diverging, non-linear patterns indicate that the relationship between aortic size and growth rate differs significantly by sex across the spectrum of initial Z-scores. Plot axes are truncated for improved visualization outlier points (n=17) with an initial Z-score >12.5 or an absolute growth rate >5 mm/year not displayed.

Overall, we observed that the GAM provided a superior fit to the best-fitting linear model (AIC= 8515 vs. 8532). Despite the overall predictive power of both models remained low, although the GAM explained a greater proportion of the variance in growth rate compared to linear models (adjusted R² 0.036 vs. 0.011). These findings remained unchanged in sensitivity analysis restricted to patients without syndromic aortopathies.

### Sub-Cohort - Continuous versus Episodic Growth (Patient-level)

While GAM analysis demonstrates how growth rate scales with baseline size at the population level, prediction intervals remained wide relative to clinically meaningful thresholds, and residual patterns show marked within-patient variability and occasional large interval-to-interval changes, features suggesting heterogeneous, time-varying dynamics that a single smooth curve cannot represent. We therefore shifted to a patient-level time-series analysis, focused on characterizing trajectories as stable, continuous, or episodic, given that clinical decisions hinge on such patient-level dynamics.

The analysis of growth patterns was conducted on the Sub-Cohort of 1055 patients who had at least four serial imaging studies. The baseline characteristics of the overall study population are detailed in **Table S5**. As described in the Methods, patients were first stratified based on their total growth over the entire surveillance period, calculated via linear regression. This yielded 760 patients classified as “Stable” (total growth <2 mm) and 295 patients as those with “Growth” (total growth ≥2 mm). Compared to the Stable group, the Growth group was significantly younger (56.2 vs. 61.9, p<0.001), had smaller baseline aortic diameters (baseline Z-score 1.5 vs. 1.9, p=0.002), higher rates of Marfan syndrome, and were more likely to undergo prophylactic repair during a longer follow-up period (**Table S6**). Notably, among the Growth group (n=295), 172 (58.3%) had a non-dilated baseline aorta (Z-score <2).

To investigate the temporal nature of growth, all patients were sub-stratified based on the presence of an episodic growth event (a diameter increase of ≥2.0 mm within a single or adjacent intervals), yielding four distinct patterns. The primary finding emerged from the Growth group (n=295), where expansion was overwhelmingly episodic: 238 patients (81%) experienced their growth in discrete bursts (“Episodic Growth group”), while only 57 (19%) grew in a steady, continuous manner (“Continuous Growth group”). For context, among the 760 Stable patients, 228 (30%) had intervals that qualified as episodes despite not demonstrating ≥2 mm of total growth over the entire surveillance duration; such alternating positive and negative changes in measurements were felt to reflect measurement noise (“Stable with noise-driven episodes”). A detailed comparison of baseline characteristics across these four patterns is presented in **Table S7**.

The distinct longitudinal trajectories of the Stable, Continuous Growth, and Discontinuous Growth groups are visualized in the lasagna plots in **Figure 5**, with a comparison of the two stable patterns provided in **Figure S8**. The Stable group shows uniformly low growth rates, while the Episodic Growth exhibit a background of stability punctuated by high-rate growth events, a pattern distinct from the persistent, low-level growth of the Continuous Growth.

**Figure 5.**
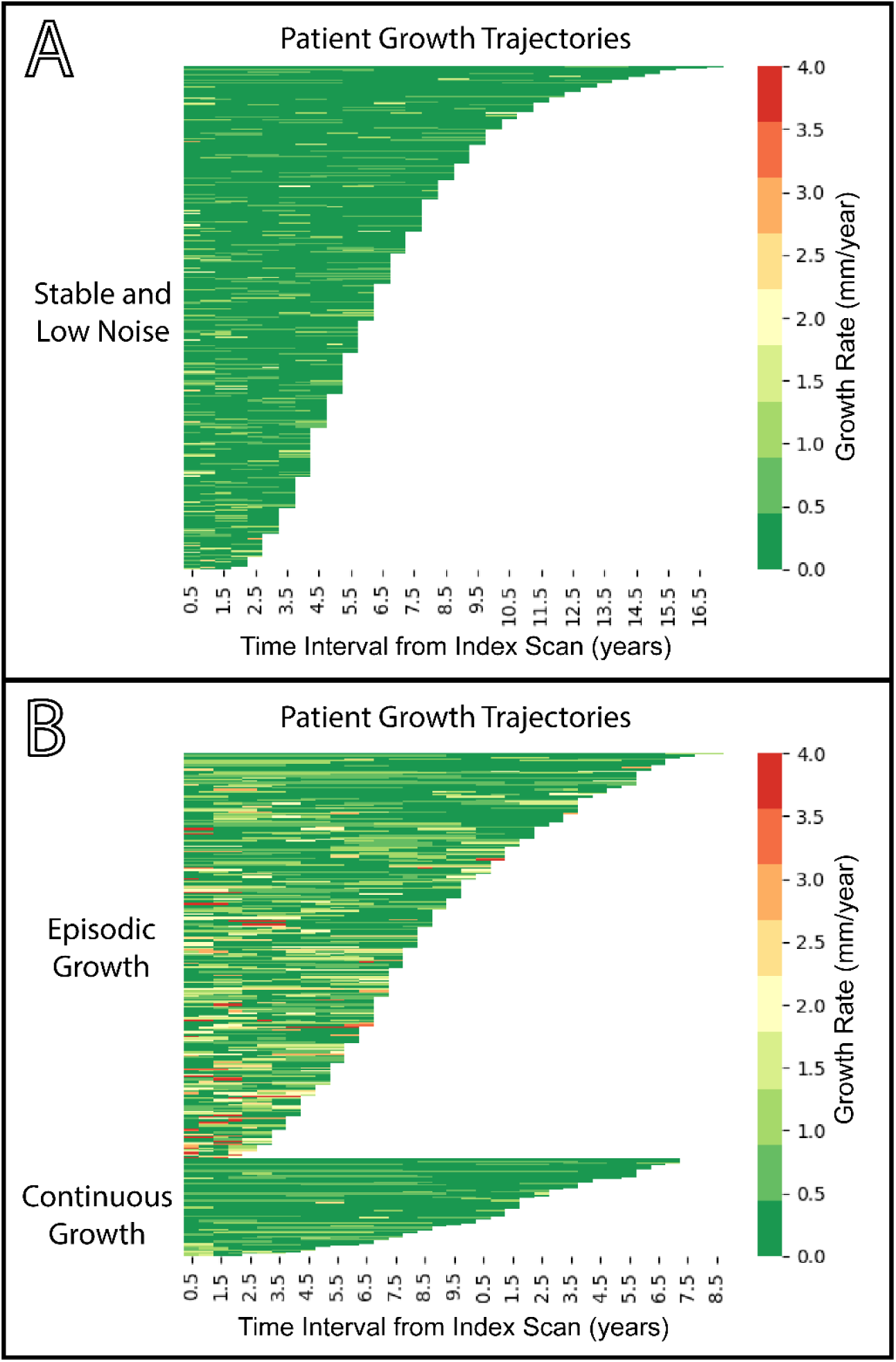
Lasagna Plots Visualizing Aortic Growth Trajectories Across Groups. Each row represents an individual patient, ordered by total follow-up duration. Each colored cell represents a single imaging interval, with the color corresponding to the annualized aortic growth rate (mm/year) during that period. **A:** Stable pattern (n=532) was characterized by a predominance of slow growth (<0.5 mm/year), but total overall growth magnitude below limits of measurable change (< 2mm). **B**: The Discontinuous Growth pattern (n=238) displays a background of slow growth intervals punctuated by sporadic, high growth intervals (>1.5 mm/year), indicating episodic growth. Conversely, the Continuous Growth pattern (n=57) shows a mixture of slow and moderate growth up to rates of approximately 1 mm/year, reflecting relatively consistent growth over surveillance.

Imaging modality was largely consistent over time. At the patient level, the percentage of individuals who experienced at least one change in imaging modality was similar between the stable and growth groups (10.2% vs. 12.5%, p=0.346). The frequency of discordant modality intervals (e.g., CT-measurement to MRI-measurement) was slightly higher in intervals with growth ≥2 mm compared to those without growth (6.0% vs. 3.9%), although this difference did not reach statistical significance (p=0.052).

## DISCUSSION

This study examined longitudinal imaging data from a large cohort of patients to better understand the natural history of ascending aortic aneurysms. Our analysis demonstrates that aortic growth is a complex process best described by two key principles. First, at the population level, the relationship between baseline aortic size and subsequent growth is not linear but follows a more complex, U-shaped curve, with this relationship being modified by well-described patient factors such as age, sex and blood pressure. Importantly, this U-shaped association revealed a group of patients in Z<0 range with mean growth rates typically encountered in aortic dilation despite “normal” aortic size. Conversely, we noted a clear inflection point towards accelerated growth in the range of Z>5, corresponding to a mean diameter of 48 mm, a size that is just below current Class 2A prophylactic surgical repair thresholds, providing further evidence to support current Class 2A guideline repair thresholds.^1,2^ Second, our patient-level analysis identified evidence that aortic growth is not a steady, monotonic – and thus predictable process – but is rather often characterized by episodic bursts of growth with intervening periods of relative stability. Taken together, our findings indicate that conventional linear-continuous growth models may not fully capture the variable activity of biologic processes that drive disease progression. Further our results support a more nuanced approach to predicting aortic growth at the patient-level that considers not just indexed size, but the important modifying effects of factors such as age and sex.

### Non-linear Growth Dynamics and the Aortic Paradox

In our population-level analysis of body size–adjusted aortic diameter versus growth rate, growth was non-linear across size strata, following a U-shaped pattern. This observation may help reconcile conflicting literature reports based on linear assumptions—particularly in cohorts dominated by non-surgical diameters (4.0–5.0 cm).^17^ Prior work has hinted at this: a large echocardiography surveillance study (n=1,414) reported a similar non-linear relationship at comparable sizes.^18^

Our study is distinctive for its size (n=3,315) and its inclusion of subjects with aortas in the non-dilated range (Z-score <0). We intentionally included those with serial ascending aortic measurements, but “normal” diameters, on the premise that growth must occur before patients reach conventional threshold definitions of dilation. Within this range, we observed accelerated growth at smaller sizes – the left arm of the U-shaped curve – which may help explain the “aortic paradox” (i.e., the disconnect between size and individual risk). Notably, a nationwide Australian cohort (n=524,994) found that nearly 60% of fatal type A dissections occurred at <4.0 cm.^10^

Consistent with this, our data suggest a subset of “normal-sized” aortas with a more aggressive phenotype: these patients were younger and more likely to have heritable thoracic aortic disease and distal aortic involvement (descending/abdominal), pointing to systemic drivers of growth. Mechanistically, this pattern may reflect an early, less-compensated phase of diseased by elastin degradation before collagen-mediated remodeling tempers expansion^19^, a decompensation-to-compensation transition analogous to heart failure, cirrhosis, and chronic kidney disease.^20-22^

Importantly, interpretation of our findings should be limited to patients with non-dilated ascending aortas already under imaging surveillance – most commonly for non-ascending aortic pathology (63.1%) or known HTAD (5.4%) – and therefore describe a higher-risk surveillance cohort rather than the general population.

The remainder of the U-shaped curve may reflect subsequent phases of remodeling. While individual variability is higher, the nadir (approximately Z-score +2 to +5) likely represents a trend towards a more compensated state in which active biological processes – such as vascular smooth muscle cell phenotypic switching – reinforce the wall and slow dilation.^23^ This dynamic balance between degradation and repair may also underlie mixed findings regarding the predictive value of aortic stiffness (ref). Finally, the upswing in growth rate at Z-score >5 (median diameter ∼48-49 mm) suggests exhaustion of compensatory mechanisms and a transition to rapid decompensation (ref), consistent with prior reports of a “hinge point” above which the risks of dissection and rupture increase sharply.^24^

### Modifiers of the Non-Linear Size-Growth Relationship

While the U-shaped curve describes the population-level trend, this relationship was significantly modified by several key clinical factors. Our analysis revealed that larger aneurysms were associated with faster growth in younger patients, a finding consistent with the more aggressive nature of genetically mediated aortopathies that manifest earlier in life.^25^ Blood pressure also demonstrated a complex, non-linear interaction with aortic size. Notably, the effects of systolic blood pressure and pulse pressure were dominant at opposite ends of the size spectrum. Higher systolic blood pressure was associated with accelerated growth primarily in non-dilated aortas, suggesting it may be a key factor in initiating early vessel wall damage.^26^ Conversely, higher pulse pressure was most impactful in severely dilated aortas, possibly indicating that increased pulsatile stress exacerbates damage in an already stiff, collagen-remodeled aortic wall.^27,28^ We also observed a significant interaction between sex and aortic size. At larger diameters, women exhibited substantially faster growth than men. This finding mirrors reports on sporadic TAA, where female sex has been frequently linked to more aggressive disease progression.^29-31^

### Episodic Growth and Punctuated Equilibrium

Our second major finding is that aortic growth is frequently a discontinuous, or episodic, process. This "staccato" growth pattern, previously suggested in abdominal aortic aneurysm (AAA) surveillance studies, was prominent in our cohort; 81% of patients with measurable growth demonstrated changes in aortic measurements suggesting discrete episodes rather than as a steady, continuous expansion.^11^ Such an episodic/discontinuous pattern is biologically plausible and can be conceptualized through the lens of punctuated equilibrium, a central theory in evolutional biology that has been more recently applied in medicine, where long periods of relative stability are interrupted by short bursts of rapid change, a dynamic also observed in some models of tumor evolution.^32,33^

Such episodes may be initiated by clinical triggers that disrupt the aortic wall’s homeostasis, such as periods of uncontrolled hypertension or systemic inflammation. From an episodic growth perspective, poor BP control intermittently elevates wall stress and perturbs vasa-vasorum perfusion, provoking ROS/protease-driven ECM injury and perhaps short-erm growth spurts followed by compensatory, collagen-heavy remodeling.^23,34-36^ The potential role of transient inflammation is supported by recent findings from Dolapoglu et al., who showed that elevated inflammatory markers, such as C-reactive protein, were associated with faster growth.^37^ Biologically, these triggers may induce vascular smooth muscle cell (VSMC) phenotypic switching, altering the balance between matrix synthesis and degradation and leading to periods of wall instability and net growth.^23^

### Limitations

Our study has several limitations. First, being a retrospective, single-center analysis, misclassification of patient data or scan indications is possible. Second, because the cohort comprises patients already undergoing serial CTA for known vascular disease, selection bias limits generalizability to a surveillance population rather than the broader community; nonetheless, this higher-risk subgroup is both currently accessible and most likely to benefit from refined risk stratification. Third, clinical diameter measurements carry inherent noise–even with standardized 3D lab protocols– which can produce spurious growth episodes or obscure true ones. We mitigated this by testing alternative episode definitions and using averaging methods less sensitive to single-interval changes, but residual misclassification is likely. Fourth, our definition of growth events relied on an absolute threshold of ≥2 mm, which does not account for differences in surveillance interval length. Clinically, however, such an absolute cutoff is commonly used clinically as it reflects the margin needed to overcome measurement noise. Importantly, we did not observe an increased frequency of growth events in patients with longer follow-up intervals; in fact, longer intervals were associated with greater stability. Fifth, a retrospective design also constrains causal inference; for example, the effect of hypertension is difficult to isolate given high rates of treatment with beta-blockers and other antihypertensives. Sixth, at the population level, our GAM uses annualized growth rates, an averaging approach that necessarily smooths the episodic dynamics observed in the sub-cohort. Accordingly, these results should be considered hypothesis-generating and motivate prospective trajectory studies using emerging, sub-millimeter measurement techniques to reduce noise.^38,39^ Despite these limitations, the large sample and flexible modeling approach reveal patterns that may be underappreciated with conventional linear approaches.

## CONCLUSION

In conclusion, our findings demonstrate that ascending aortic growth is not a simple linear process but is defined by complex, non-linear size-growth dynamics. Furthermore, our patient-level analysis strongly suggests that this growth is also episodic/ punctuated. While the characterization of this episodic pattern is more hypothesis-generating due to inherent measurement variability, acknowledging this dual reality—that growth is neither linear nor continuous— is a critical step toward studying more accurately understanding the complex and variable natural history of this disease.

## Data Availability

Data for this study will be made available upon reasonable request and within the limitations of restrictions of sharing protected health information.

## Abbreviations

ACTA2: actin alpha-2, smooth muscle (gene)
AsAo: ascending aorta
BP: blood pressure
GAM: Generalized Additive Model
IQR: interquartile range
MYH11: myosin heavy chain 11 (gene)
MYLK: myosin light chain kinase (gene)
PP: pulse pressure

## Funding statement

**Nicholas S. Burris**: National Institutes of Health (R01HL170059).

## Disclosures

- Dr. Burris is entitled to royalties related to licensure of intellectual property of the vascular deformation mapping technology to Imbio Inc.; coinventor of vascular deformation mapping technique (U.S. patent 10,896,507 [techniques of deformation analysis for quantification of vascular enlargement]). Dr. Burris is supported in part by grants from the National Institute of Heart, Lung and Blood Insitute (R01HL170059-02). He is also supported by the David A. Bluemke MD PhD Professorship in Radiology.
- Dr. Murthy owns stock or stock options in General Electric, Cardinal Health, Ionetix, Boston Scientific, Merck, Eli Lilly, Johnson and Johnson, and Pfizer. He has received research grants and consulting fees from Siemens Medical Imaging and consulting fees from Jubilant Draximage and Credence Management Solutions. He has served on medical advisory boards for Ionetix. Dr. Murthy is supported in part by grants from the National Institute of Diabetes, Digestive, and Kidney Diseases (U01DK123013-03); and National Institute on Aging (R01 AG059729). He is also supported by the Melvyn Rubenfire Professorship in Preventive Cardiology.
- Otherwise, all authors declare freedom of investigation, and no conflict of interest is related to the contents of this manuscript.

## Notes

### Funding Statement

No external funding was received for this work.

### Author Declarations

This retrospective analysis was performed as part of an Institutional Review Board approved study [University of Michigan Medical Schools' IRB (IRBMed), HUM00133798], and informed consent was waived.

